# Assessing the Value of ChatGPT for Clinical Decision Support Optimization

**DOI:** 10.1101/2023.02.21.23286254

**Authors:** Siru Liu, Aileen P. Wright, Barron L. Patterson, Jonathan P. Wanderer, Robert W. Turer, Scott D. Nelson, Allison B. McCoy, Dean F. Sittig, Adam Wright

**Affiliations:** Department of Biomedical Informatics, Vanderbilt University Medical Center, Nashville, TN, USA; Department of Medicine, Vanderbilt University Medical Center, Nashville, TN, USA; Department of Pediatrics, Vanderbilt University Medical Center, Nashville, Tennessee, USA; Department of Anesthesiology, Vanderbilt University Medical Center, Nashville, Tennessee, USA; Department of Emergency Medicine, University of Texas Southwestern Medical Center, Dallas, TX, USA; Clinical Informatics Center, University of Texas Southwestern Medical Center, Dallas, TX, USA; School of Biomedical Informatics, University of Texas Health Science Center, Houston, Texas, USA

**Keywords:** artificial intelligence, clinical decision support, large language model

## Abstract

**Objective:** To determine if ChatGPT can generate useful suggestions for improving clinical decision support (CDS) logic and to assess noninferiority compared to human-generated suggestions.

**Methods:** We supplied summaries of CDS logic to ChatGPT, an artificial intelligence (AI) tool for question answering that uses a large language model, and asked it to generate suggestions. We asked human clinician reviewers to review the AI-generated suggestions as well as human-generated suggestions for improving the same CDS alerts, and rate the suggestions for their usefulness, acceptance, relevance, understanding, workflow, bias, inversion, and redundancy.

**Results:** Five clinicians analyzed 36 AI-generated suggestions and 29 human-generated suggestions for 7 alerts. Of the 20 suggestions that scored highest in the survey, 9 were generated by ChatGPT. The suggestions generated by AI were found to offer unique perspectives and were evaluated as highly understandable and relevant, with moderate usefulness, low acceptance, bias, inversion, redundancy.

**Conclusion:** AI-generated suggestions could be an important complementary part of optimizing CDS alerts, can identify potential improvements to alert logic and support their implementation, and may even be able to assist experts in formulating their own suggestions for CDS improvement. ChatGPT shows great potential for using large language models and reinforcement learning from human feedback to improve CDS alert logic and potentially other medical areas involving complex, clinical logic, a key step in the development of an advanced learning health system.

## INTRODUCTION

Clinical decision support (CDS) provides information and recommendations to healthcare professionals and patients at the point of care.[1] As electronic health record (EHR) adoption has increased, in part due to more than $34 billion of government spending,[2] the use of CDS has also increased. Rule-based CDS alerts that deliver patient- and task-specific recommendations are a required part of all certified EHRs.[3] Such CDS alerts can improve clinical practice,[1,4] standardize care to close quality gaps,[5] and address racial and ethnic disparities.[6] For example, a review of cardiovascular disease studies found that CDS alerts increased guideline-recommended testing and examinations by 9.6%-45.6% in Black and Hispanic populations.[7] A classic CDS design framework attempts to optimize CDS utility by ensuring that they are relevant to the right patient, to the right health professional, at the right time in the workflow, in the right intervention format, and through the right channel.[8]

Despite these potential benefits, approximately 90% of alerts are overridden or ignored by clinicians with justifiable reasons (e.g. irrelevancy, poor timing, or incomplete characterization of clinical condition).[9–11] Alert fatigue arises when clinicians encounter these poorly performing alerts, which threatens patient safety.[12–14] Researchers have proposed several approaches for optimizing alerts. The first approach uses human review to optimize alert content, timing, and target audience,[15–17] which can reduce 9% to 35% of alerts with no untoward consequences.[18] For example, Vanderbilt University Medical Center (VUMC) conducted the Clickbusters program,[19] where 24 physicians and informaticians reviewed alerts at VUMC using a structured process. They eliminated 70,000 unnecessary weekly alert firings, representing a 15% decrease. However, this approach is resource-intensive, subject to cognitive bias and premature closure, and requires periodic re-assessment. Marginal effects also diminish rapidly (i.e., after all simple improvements have been identified, it takes a disproportionate effort for reviewers to identify further improvements).[20,21] Experts involved in manual reviews are often clinicians who practice in a single area or use a particular workflow. Subsequently, they may not consider improvements that are relevant to other team members with different workflows. Automated tools using simple rules or machine learning techniques for identifying problematic alerts might allow for sustainable and scalable CDS maintenance.[22,23]

Valuable insights generated by ChatGPT or other large language models could greatly support experts in refining their suggestions and enhance the specificity of alerts, ultimately addressing the issue of alert fatigue. ChatGPT, an artificial intelligence (AI) model created by OpenAI, has achieved attention for its ability to solve a wide range of natural language processing tasks and generate human-like responses. ChatGPT was built using the GPT-3.5 large language model and fine-tuned for general tasks using human feedback and supervised and reinforcement learning.[24] ChatGPT was launched on November 30, 2022, and has gained widespread attention among the medical community due to its potential to semi-autonomously perform tasks such as answering sample questions from United States Medical Licensing Exam (USMLE) and generating simplified radiology reports for patient consumption.[25,26] As a novel AI algorithm, the goal of the large language model is to predict the next sequences of words based on the previous context. To achieve this goal, large language models are often pre-trained with large text corpora. For example, the large language model used in ChatGPT, GPT3.5, was trained with 175 billion parameters, with a dataset including CommonCrawl and WebText (web page data until 2021), two Internet-based book corpora, and English-language Wikipedia.[27] After generating the large language model, OpenAI sampled prompts and collected related demonstration data from humans. Then, they used this dataset for supervised learning to fine-tune the GPT3.5 language model. In a second step, they asked annotators to rank the model outputs based on quality, which was used as a reward function to further fine-tune the supervised learning model to maximize the reward.

To address current challenges in optimizing CDS alerts, we proposed that chatGPT-based CDS alert tuning might enable fast and cost-effective analysis of a high volume of alerts. The objectives for this work were to determine if ChatGPT can generate useful suggestions for improving clinical decision support logic and to assess noninferiority compared to human-generated suggestions. Our goal was not to show that ChatGPT suggestions are superior to human suggestions but, rather, to show that the ChatGPT suggestions may enhance traditional techniques for CDS maintenance and optimization. This is consistent with the fundamental theorem of medical informatics[28], where the goal is not to create computer systems that are superior to human, but rather to create systems that augment human intelligence, such that the human and computer together perform better than the human alone.

## METHODS

### Setting

This project was conducted at VUMC, a large integrated delivery system in the Southeastern United States, which uses Epic (Epic Systems Co., Verona, WI) as its EHR. VUMC has more than 80 certified Physician Builders, who are trained and certified to develop and maintain CDS, have experience in using CDS tools, and are willing to participate in EHR-related projects within the organization. We previously conducted an alert optimization program – Clickbusters – involving a 10-step process of reviewing alert related data and clinical evidence, identifying possible improvements, discussing improvements with stakeholders, making changes in the test environment, testing, and evaluating.[19] The entire process was documented and archived.

### Human-Generated Suggestions

In this study, we analyzed 7 alerts (BestPractice Advisories) from the Epic EHR at VUMC. These alerts, described in Table 1, were selected from the previously described Clickbusters program, because they had previously been reviewed for suggested improvements by clinical informaticians.[19] During the review process, the alert logic and human suggestions were documented.

**Table 1.** Selected alerts and descriptions.

| Alert Title | Description |
| --- | --- |
| a1: Immunocompromised and Live Virus Immunization | To prevent ordering a live virus vaccine for patients who are immunosuppressed. |
| a2: Anesthesia Postoperative Nausea and Vomiting | To identify patients who have risk factors for postoperative nausea and vomiting (PONV). |
| a3: Pediatrics Bronchiolitis Patients with Inappropriate Order | To identify potentially inappropriate use of albuterol or chest x-rays in children with bronchiolitis |
| a4: Artificial Tears Frequency > 6/day | To identify patients who have been prescribed artificial tears with a frequency exceeding 6 administrations per day. |
| a5: IP Allergy Documentation | To identify inpatients over 8 weeks old who have documented allergies, but have not had their allergy list reviewed |
| a6: RX NSAID/Pregnancy | To discourage ordering non-steroidal anti-inflammatory drugs (NSAIDs) in pregnant patients. |
| a7: Warfarin No INR | To notify pharmacists upon order verification of warfarin if the patient does not have an international normalized ratio (INR) result in the past 7 days. |

### AI-Generated Suggestions

To generate potential improvements to the alerts automatically, we selected the ChatGPT chatbot. We transformed the documented alert logic into ChatGPT prompts in the following format: “I have a clinical decision support alert about [alert description]. [statement of guideline or standard, if available]. Current inclusion and exclusion criteria are listed below. Are there any additional exclusions that should be added? [Inclusion and exclusion criteria of the alert].” In addition, for groups of specific medications and diagnoses, we used additional prompts, such as “what other immunosuppressant medications should be added?” An example of using ChatGPT to generate suggestions to improve an alert is listed in Figure 1. All prompts used in this study are listed in Appendix 1.

**Figure 1.**
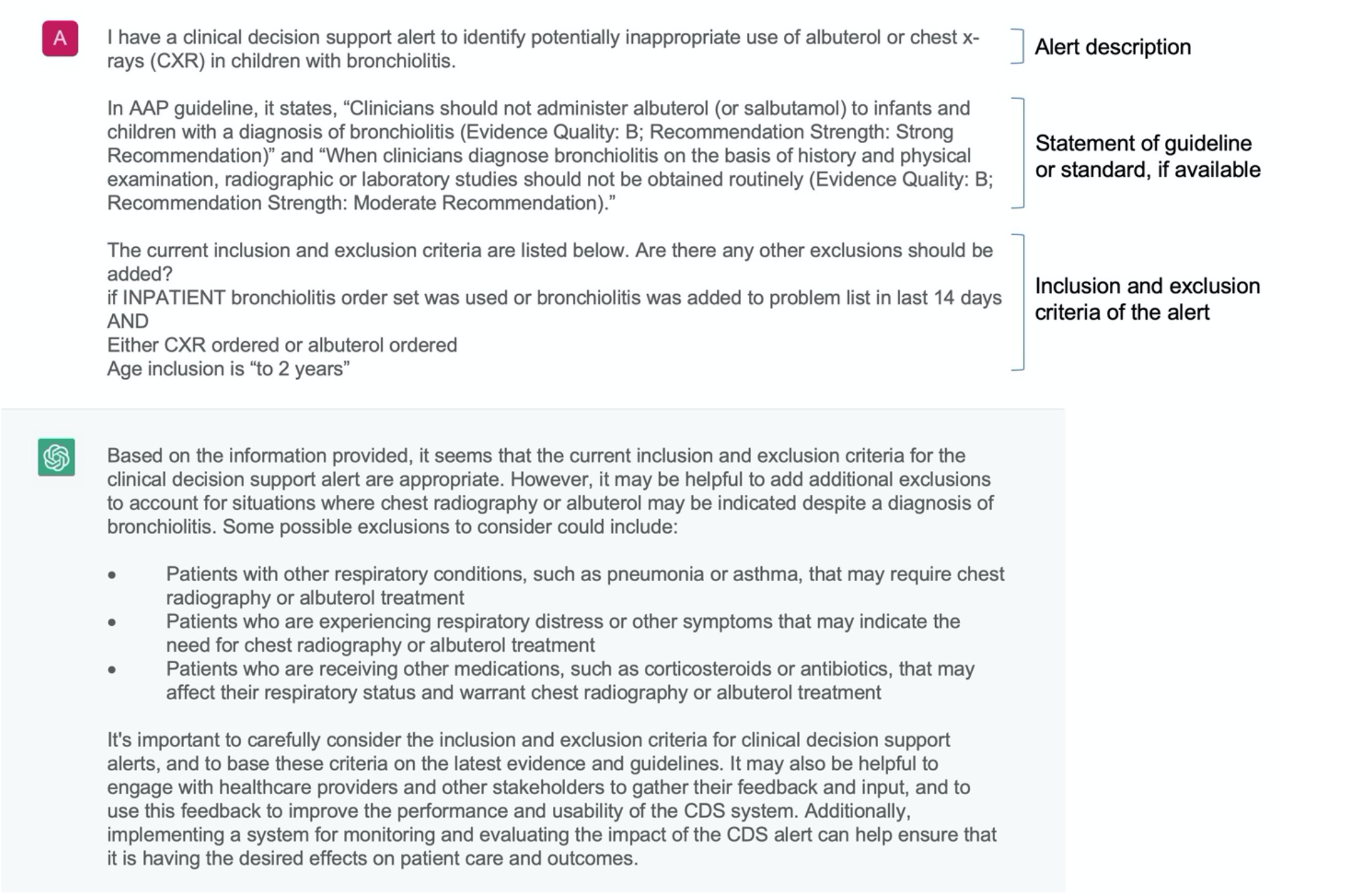
An example of using ChatGPT to generate suggestions to improve an alert.

### Expert Review of Suggestions

We mixed AI-generated suggestions with the suggestions previously generated by clinical informaticians (4 physicians and 1 pharmacist) during the Clickbusters program and grouped them by alert.[19] Within each alert, we randomized the order of the AI- and human-generated suggestions. For the human-generated suggestions, we reformatted them if they included specific alert identifiers, since humans often included these identifiers but ChatGPT never did (because it did not have access to record IDs). For example, if a human suggested something like “Exclude patients with medications in grouper [1234]” and “grouper [1234]” corresponded to “immunosuppressant medications” we transformed the human-generated suggestion to “Exclude patients with immunosuppressant medications.”

We created a semi-structured questionnaire using REDCap.[29] For each alert, we listed 1) the alert description, 2) the logic of the alert, and 3) a link to more detailed information (e.g., a screenshot of the alert). The questionnaire content was piloted within the research group (AW, AW, and AM) to improve its structure. Participants were a convenience sample of physicians and pharmacists with formal training in informatics and professional experience optimizing CDS tools. Participants were recruited from VUMC and UT Southwestern Medical Center. Each participant was assigned a unique number to ensure anonymity. Each suggestion was independently rated on a 5-point Likert scale (1-strongly disagree, 5-strongly agree) from eight perspectives: 1) **Understanding**: I understand this suggestion. 2) **Relevance**: This suggestion includes relevant concepts. 3) **Usefulness**: This suggestion contains concepts that will be useful for improving the alert. 4) **Acceptance**: I can accept this suggestion without edits. 5) **Workflow**: Based on this suggestion, I will recommend a change to a clinical workflow/process outside of this alert. 6) **Redundancy**: This suggestion is redundant with the existing alert logic. 7) **Inversion**: This suggestion is inverted (e.g., the suggested exclusion should be an inclusion). 8) **Bias**: This suggestion may contribute to bias. We also included a text box for each suggestion where participants could provide additional comments. For example, a common problem with AI-generated text is the tendency of language models to make up information, a phenomenon known as “hallucination.”[30]

### Evaluation

We calculated the mean (standard deviation) for each item for each suggestion and generated box plots to show median and interquartile range, etc. In the overall score calculation, for Redundancy, Inversion, and Bias items, we used reversed scores, i.e. (1 -strongly agree, 5 -strongly disagree). The overall score was the average of the three reversed scores and the five scores for the remaining items. The calculation process was then repeated at the alert level. We performed a nonparametric Mann-Whitney Wilcoxon Test to compare expert ratings of model-generated suggestions and manual review-generated suggestions.[31] In the alert-level analysis, we conducted Kruskal-Wallis H-tests to compare median values for each item. Statistical significance was set at *P*<0.01. In addition, we calculated the intra-class correlation coefficient (ICC) to evaluate interrater reliability.[32] ICC estimates and 95% confidence intervals were reported based on a 2-way mixed-effects model (mean of k raters type and consistency definition). For ICC estimates, below 0.5 represents low reliability; 0.5 to 0.74 represents moderate reliability; 0.75 to 0.9 represents good reliability; and greater than 0.9 represents excellent reliability.[33] Statistical analyses were conducted in Python3.6. We analyzed free text responses, grouped them using thematic analysis and reported summarized themes. We also reported descriptive statistics of participants’ characteristics (e.g., clinical service, roles, and years of experience with CDS).

## RESULTS

Five CDS experts trained in internal medicine, anesthesiology, pharmacy, emergency medicine, and pediatrics participated in the survey. The average values of clinical experience and EHR experience were 13.75 years and 16.25 years, respectively. The characteristics of participants in the survey are listed in Table 2. The ICC value was 0.86, with a 95% confidence interval ranging from 0.83 to 0.87, indicating good reliability.

**Table 2.** Characteristic of 5 CDS experts participating in the survey.

|  |  |
| --- | --- |
| **Gender** |  |
| Male | 4 |
| Female | 1 |
| **Clinical Specialty** |  |
| Internal medicine | 1 |
| Anesthesiology | 1 |
| Pharmacy | 1 |
| Emergency Medicine | 1 |
| Pediatrics | 1 |
| **Clinical role** |  |
| Physician | 4 |
| Pharmacist | 1 |
| **Years of clinical experience** | 13.75 |
| **Years of EHR experience** | 16.25 |

### Examples of AI-Generated Suggestions and Human-Generated Suggestions

The final questionnaire included 36 AI-generated suggestions and 29 human-generated suggestions for 7 alerts. Of the 20 suggestions that scored highest in the survey, 9 were generated by ChatGPT. These 20 suggestions and their scores for acceptance, relevance, understanding, and usefulness are presented in Table 3.

**Table 3.** Top 20 suggestions and their ratings for acceptance, relevance, understanding, and usefulness.

|  | Suggestion | Alert | Acceptance | Relevance | Understanding | Usefulness |
| --- | --- | --- | --- | --- | --- | --- |
| Human | Scope of roles should include fellow. | a2 | 4.8±0.5 | 4.8±0.5 | 4.8±0.5 | 4.8±0.5 |
| Human | Pregnant patients. | a1 | 3.0±1.0 | 4.8±0.5 | 4.4±0.9 | 4.8±0.5 |
| AI | Some examples of additional medications or treatments that may have immunosuppressive effects include certain cancer treatments, such as chemotherapy or radiation therapy, as well as certain medications used to treat autoimmune disorders, such as rituximab or infliximab. | a1 | 2.4±1.1 | 4.8±0.5 | 4.8±0.5 | 4.6±0.6 |
| AI | Exclude: Patient is currently receiving radiation or chemotherapy treatment. | a1 | 2.6±1.5 | 4.4±0.6 | 3.8±1.6 | 4.5±0.6 |
| Human | Add an active chemotherapy criterion. | a1 | 2.4±0.9 | 4.8±0.5 | 4.8±0.5 | 4.4±0.6 |
| AI | Add biologic agents, such as adalimumab, etanerfigut [sic]*, and golimumab, which are used to treat autoimmune disorders. | a1 | 2.2±1.1 | 4.8±0.5 | 4.6±0.6 | 4.4±0.6 |
| AI | Exclude: Patients who have recently undergone bone marrow transplant or solid organ transplant. | a1 | 2.2±1.3 | 4.6±0.6 | 4.0±1.7 | 4.3±0.5 |
| Human | Make the list of immunosuppression medications more complete. | a1 | 2.4±1.1 | 4.6±0.6 | 4.6±0.6 | 4.2±0.8 |
| Human | Exclude NICU and newborn departments. | a5 | 3.4±1.3 | 4.2±0.5 | 4.4±0.6 | 4.2±0.5 |
| AI | Exclude patients with other respiratory conditions, such as pneumonia or asthma, that may require chest radiography or albuterol treatment. | a3 | 2.6±0.9 | 4.6±0.6 | 4.8±0.5 | 4.0±1.2 |
| AI | Add medications used to treat transplant rejection, such as basiliximab, daclizumab, and muromonab-CD3. | a1 | 2.4±1.1 | 4.4±0.6 | 4.6±0.6 | 4.0±1.2 |
| Human | Currently history of PONV looks to problem list, but it should also look to the last anesthesia preop evaluation. | a2 | 3.4±0.9 | 4.8±0.5 | 4.4±0.9 | 4.0±1.0 |
| AI | Exclude: Currently receiving immunosuppressive therapy or who have recently completed such therapy. | a1 | 1.8±1.1 | 4.20±0.5 | 3.6±1.5 | 4.0±0.8 |
| Human | Add additional eye drops. | a4 | 2.4±0.9 | 4.2±0.5 | 4.4±0.6 | 4.0±0.7 |
| Human | Add a 3rd OR statement if the ED bronchiolitis panel was used during this encounter. | a3 | 3.2±1.6 | 3.8±1.6 | 3.8±1.6 | 3.8±1.6 |
| Human | Restrict inpatient options to only formulary. | a4 | 3.2±0.8 | 3.8±1.1 | 4.0±1.2 | 3.8±1.1 |
| Human | Add inpatient orders. | a4 | 3.6±0.9 | 3.8±1.1 | 3.8±1.1 | 3.8±1.1 |
| AI | Exclude patients who are receiving palliative care or end-of-life care, as these patients may require chest radiography or albuterol treatment for symptom management. | a3 | 3.2±1.6 | 3.6±1.7 | 4.4±0.9 | 3.6±1.7 |
| Human | Add patients with transplant on problem list. | a1 | 2.6±1.3 | 4.6±0.6 | 4.6±0.6 | 3.6±1.5 |
| AI | Patients who have had previous PONV episodes, as they may be at higher risk for developing PONV again. | a2 | 2.0±1.0 | 4.6±0.6 | 4.6±0.6 | 3.6±1.1 |

\*. Etanerfigut is either a misspelling, which is hard to imagine from a computer, or a nonexistent medication hallucinated by ChatGPT.

### Results of Expert Review of AI-generated Suggestions and Human-generated Suggestions

Out of the 36 AI-generated suggestions, 27 (75%) achieved an overall score of 3 or higher, with a maximum score of 4.0±0.2 and a minimum score of 2.8±0.4. The mean score was 3.3±0.5. These AI-generated suggestions provided additional immunosuppressive medications and treatments and excluded additional patients (e.g., patients with other respiratory conditions that may require chest radiography or albuterol treatment, and patients in palliative care). On average, the scores of AI-generated suggestions for relevance and understanding were rated as “agree”, usefulness as “neither agree nor disagree”, bias as “strongly disagree”, and workflow, inverted, and redundancy as “disagree”. Figure 2 shows two stacked bar charts of the scores for each item of the AI-generated suggestions (Figure 2a) and human suggestions (Figure 2b). In Figure 2a, AI-generated suggestions had high understanding and relevance, which were similar with human-generated suggestions. Although human suggestions were more likely to be accepted, for less important features, workflow, bias, and backwards, in general, they were rated similarly by experts.

**Figure 2.**
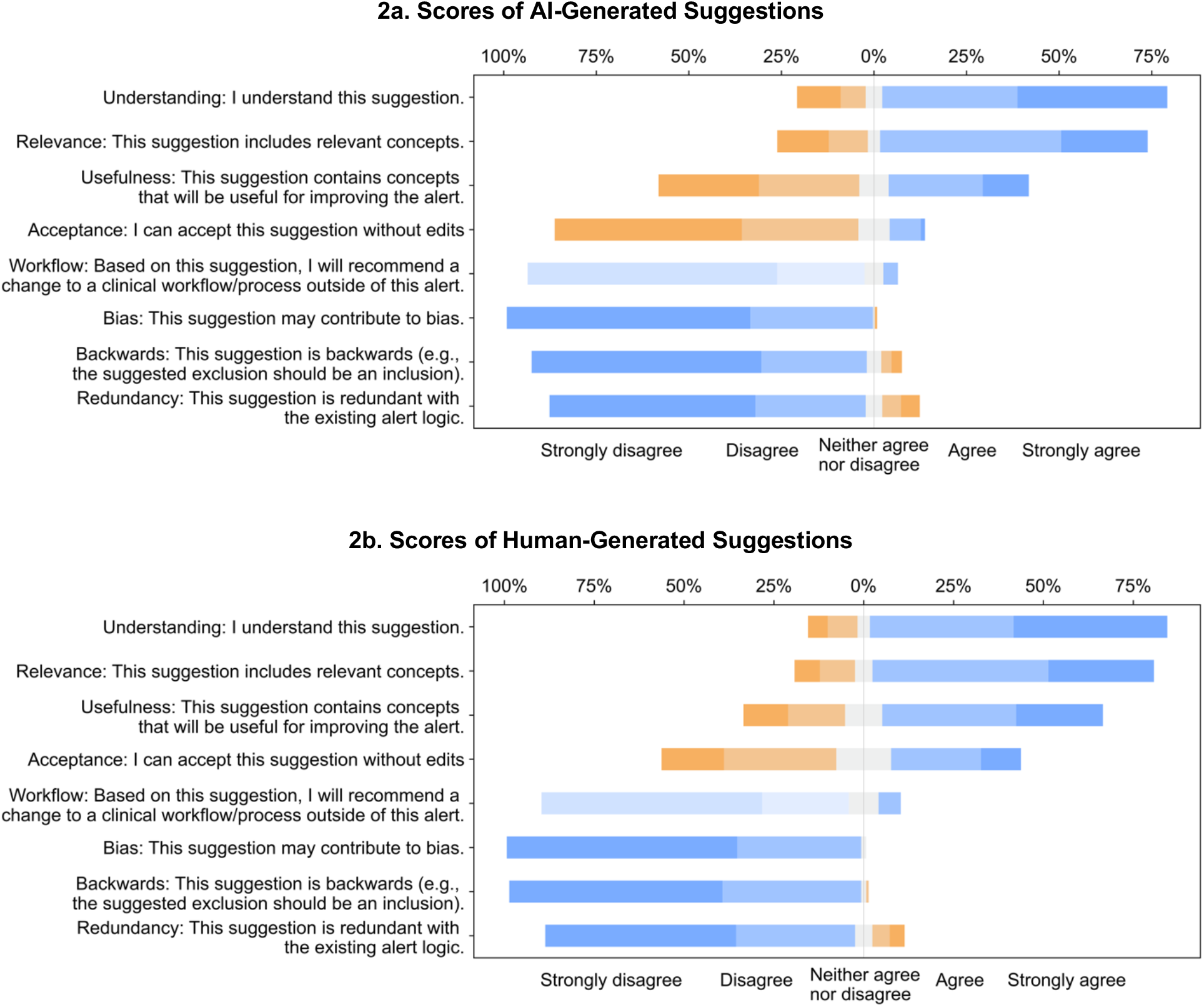
Stacked bar charts of the scores of each item (understanding, relevance, usefulness, acceptance, workflow, bias, inversion, redundancy) for AI-generated suggestions (2a) and manual review-generated suggestions (2b).

AI-generated suggestions achieved high scores in understanding and relevance and did not differ significantly from human-generated suggestions. In addition, AI-generated suggestions did not show significant differences in terms of bias, inversion, redundancy, or workflow compared to human-generated suggestions. However, AI-generated suggestions received lower scores for usefulness (AI: 2.7±1.4, human: 3.5±1.3, P<0.001) and acceptance (AI: 1.8±1, human: 2.8±1.3, P<0.001). The overall scores were human: 3.6±0.6 and AI: 3.3±0.5 (P<0.001). Values for mean and standard deviation for each item are presented in Table 4. Boxplots for each item are in Appendix 2.

**Table 4.** Means and standard deviations (std) for survey items using a 5-point Likert scale (1-strongly disagree, 5-strongly agree).

|  | AI-Generated Suggestions<br>(mean±std) | Human-Generated Suggestions<br>(mean±std) | P |
| --- | --- | --- | --- |
| **Understanding:** I understand this suggestion. | 3.9±1.3 | 4.1±1.1 | 0.3 |
| **Relevance:** This suggestion includes relevant concepts. | 3.6±1.3 | 3.8±1.2 | 0.09 |
| **Usefulness:** This suggestion contains concepts that will be useful for improving the alert. | 2.7±1.4 | 3.5±1.3 | <0.001 |
| **Acceptance:** I can accept this suggestion without edits. | 1.8±1 | 2.8±1.3 | <0.001 |
| **Workflow:** Based on this suggestion, I will recommend a change to a clinical workflow/process outside of this alert. | 1.5±0.8 | 1.6±0.9 | 0.2 |
| **Bias:** This suggestion may contribute to bias. | 1.4±0.6 | 1.4±0.5 | 0.8 |
| **Inversion:** This suggestion is inverted (e.g., the suggested exclusion should be an inclusion.) | 1.6±0.9 | 1.4±0.6 | 0.9 |
| **Redundancy:** This suggestion is redundant with the existing alert logic. | 1.7±1.1 | 1.7±1 | 0.8 |
| **Overall** | 3.3±0.5 | 3.6±0.6 | <0.001 |

We further compared the scores of each item for AI-generated suggestions grouped by alert. The results showed significant variations in the scores for usefulness, acceptance, and relevance between alerts. On the other hand, the scores for understanding, workflow, bias, inversion, and redundancy did not vary significantly between the alerts.

According to the results, the AI-generated suggestions were found to have high levels of understanding (3.9±1.3) and relevance (3.6±1.3), both of which tended to “agree.” All AI-generated suggestions scored 3 or higher on the understanding item, with 29 (80.6%) of them scoring ≥3 on relevance. For example, the AI-generated suggestion for including patients with a history of PONV in the “post-anesthesia nausea and vomiting” alert ranked third among all suggestions for both understanding and relevance. Similarly, excluding patients who have recently undergone bone marrow or solid organ transplantation from the “immunocompromised and live virus immunity” alert also ranked third for relevance.

The AI-generated suggestions had a moderate level of usefulness with a mean score of 2.7±1.4. Nearly half of the AI-generated suggestions (15; 41.7%) received a score of 3 or higher on the usefulness item. Additionally, besides the suggestions previously mentioned, two other AI-generated suggestions for the “Immunocompromised and Live Virus Immunization” alert, regarding the exclusion of patients currently receiving radiation or chemotherapy treatment and the exclusion of patients currently receiving immunosuppressive therapy or who have recently completed such therapy, ranked 2^nd^ and 5^th^ in terms of usefulness, respectively. The mean score for the acceptance item was 1.8±1, and two AI-generated suggestions (5.6%) receiving a score of 3 or higher, indicating they could be accepted without changes. These suggestions were 1) the exclusion of patients who are receiving palliative care or end-of-life care for the “Peds Bronchiolitis Patients with Inappropriate Order” alert and 2) the exclusion of patients with a pending INR test for the “Warfarin No INR” alert. It was unlikely to change workflow based on AI-generated or human-generated suggestions. Bias and redundancy were also low.

### Qualitative Analysis of Comments on AI-Generated Suggestions

#### Lack of knowledge management and implementation

The lack of knowledge management and implementation understanding was a common barrier to the acceptance of AI-Generated suggestions. For example, regarding the “Immunocompromised and Live Virus Immunization” alert, the AI-generated suggestion to “Exclude patients who are currently receiving immunosuppressive therapy or who have recently completed such therapy,” was commented on by experts that they liked the idea of excluding such patients for this alert, but they noted that it would require additional work to identify appropriate value sets and other specific implementation details in the EHR, before it could be included. Additional informatics work will be needed to operationalize these suggestions and implement them in the EHR.

#### Hallucination

In ChatGPT’s “Immunocompromised and Live Virus Immunization” alert, one of the AI-generated suggestions was to “add biologic agents, such as adalimumab, etanerfigut [sic], and golimumab, which are used to treat autoimmune disorders.” Experts pointed out that “etanerfigut” was not a medication, or perhaps it was a misspelling of “etanercept”.

#### Partially correct information

An expert commented that the AI-generated suggestion “Exclude: Patient is currently receiving radiation or chemotherapy treatment” for the “Immunocompromised and Live Virus Immunization” alert should only include chemotherapy.

#### Divergent opinions from experts

We also found that there was disagreement among experts when it came to AI-generated suggestions. One of the AI-generated suggestions for the “Anesthesia Postoperative Nausea and Vomiting” alert was “patients who are taking medications that may increase the risk of PONV, such as certain antidepressants or chemotherapy drugs.” While two participants found the suggestion useful, another expert pointed out that the suggestion was potentially misleading, as antidepressants are not typically associated with PONV, and some studies have found that they may actually decrease PONV.

## DISCUSSION

In this study, we applied ChatGPT to generate suggestions to improve the logic of CDS alerts. To evaluate AI-Generated suggestions, we mixed them with human-generated suggestions and asked CDS experts to rate all suggestions for their usefulness, acceptance, relevance, understanding, workflow, bias, inversion, and redundancy. While the AI-Generated suggestions were not scored as highly as the human-generated suggestions, our findings demonstrated that AI-generated suggestions had high relevance and understanding scores, moderate usefulness scores, and low scores in ability to improve clinical workflow, bias, inversion, and redundancy. In addition, the lack of redundancy between the human- and AI-generated suggestions indicate that the AI could supplement traditional CDS optimization, as would be expected from the fundamental theorem of medical informatics.[28]

The results of this study indicate that ChatGPT could be used to automatically analyze alert logic and generate useful suggestions. Most of the AI-generated suggestions could not be accepted without modification, but they still offered valuable insights for experts to build upon. In addition, this approach enables the rapid analysis of many alerts, making it possible to scale CDS optimization efforts. Additionally, this approach could be well-integrated into the alert development process to provide AI-generated suggestions at the alert development stage.

A prototype is shown in Figure 3, where the top section displays the CDS alert being created by the user and its corresponding logic, and the bottom section displays the suggestions generated by AI. Based on the suggestions, the user could refine the alert logic to make it more specific and consider some easily overlooked aspects. It is worth noting that Epic provides “Build Inspectors” in its development interface offer suggestions to improve the alerts such as ““Remove manual follow-up actions or configure this advisory to display to users.” However, these build inspectors provide feedback on data storage formats and specific functionality within the Epic system, rather than examining the alert logic itself. AI models like ChatGPT could provide similar recommendations, but for content.

**Figure 3.**
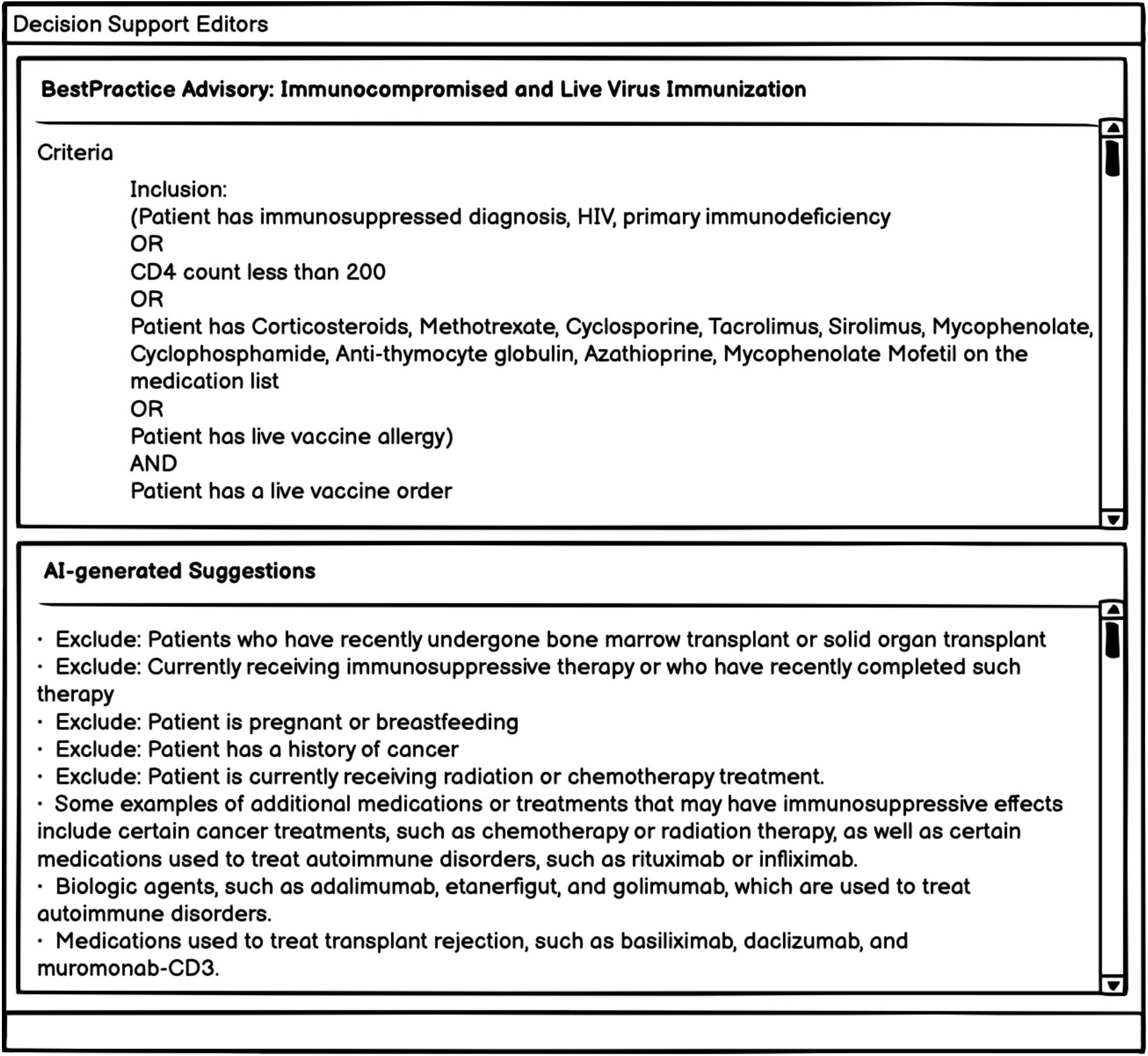
A prototype of potential implementation in EHR system – AI Decision Support Editors

In this project, we generated suggestions directly using the ChatGPT model, which was trained on a general dataset consisting of web pages, books, and Wikipedia for a variety of common use cases such as text generation, open-ended question-answers, brainstorming, chat, and rewriting in the form of conversations. Future research could therefore focus on improving language models and specifying training tasks. First, medical texts, such as clinical notes, PubMed articles from the MIMIC-III dataset, could be added to the language model. For example, using a publicly available clinical language model, GatorTron, based on 90 billion words of de-identified clinical notes from the University of Florida (UF) Health, PubMed articles, and Wikipedia, might be a good start.[34] Another example is BioGPT, a GPT-like model trained on PubMed articles.[35] In addition, UpToDate is an important source of CDS content for alert development and should also be considered for adding to the language model. Second, based on the Reinforcement Learning from Human Feedback (RLHF) framework, researchers could train a model specifically for this task on the improved language model.[36] Suggested steps include: 1) Human-generated suggestions: recruit CDS experts to generate suggestions for a number of CDS alerts (i.e., the Clickbuster process).[19] 2) Supervised fine-tuning (SFT): Use the selected alerts and human-generated suggestion dataset to fine-tune the pre-trained language model to learn a supervised strategy (SFT model) to generate suggestions for selected alerts. 3) Mimic human preferences: recruit CDS experts to rank the output of the baseline model (i.e., the generated suggestions). The generated suggestions and corresponding expert ranks are used as the reward model in reinforcement learning. 4) Proximal Policy Optimization (PPO): The reward model is used to further fine-tune and improve the SFT model to develop the final policy model.

### Limitations

This study has several limitations. First, the ChatGPT model is sensitive to the provided prompts and as a result, the AI-generated suggestions may vary based on changes in the input sentences. The prompt format used in our experiment was derived from an analysis of multiple input forms, but it is possible that there are other more effective ways to engage the ChatGPT model in this specific task. Second, we assessed the quality of AI-generated suggestions from the viewpoint of CDS experts, but the effect on clinical outcomes remains unknown. Third, ChatGPT was trained on text up to 2021 and did not include information on new drugs or clinical guidelines developed after that year. Consequently, ChatGPT is unable to provide suggestions regarding clinical guidelines and drugs that were developed after 2021.

## Data Availability

The input prompts and AI outputs are available in Appendix.

## CONCLUSION

Alert fatigue is a pressing issue. In this study, we evaluated the feasibility of using ChatGPT to generate suggestions for improving the specificity of alert logic. The suggestions generated by AI were found to offer unique perspectives and were evaluated as highly understandable and relevant, with moderate usefulness, low acceptance, bias, inversion, redundancy, and low ability to improve clinical workflow. Therefore, these AI-generated suggestions could be an important complementary part of optimizing CDS alerts, can identify potential improvements to alert logic and support their implementation, and may even be able to assist experts in formulating their own suggestions for CDS improvement. Overall, ChatGPT shows great potential for using large language models and reinforcement learning from human feedback to improve CDS alert logic and potentially other medical areas involving complex, clinical logic, a key step in the development of an advanced learning health system.

## CONTRIBUTORSHIP STATEMENT

SL applied chatGPT to provide AI-generated suggestions and conducted survey development, data extraction, statistical analysis, and drafting the work. AM, Adam W, and DS helped to design experiments AM, Adam W, and Aileen W participated in the pilot test of the survey. Aileen W, BP, JW, RT, and SN participated in the survey. SL, Adam W, AM, DS, Aileen W, BP, JW, RT, and SN revise the drafted manuscript. All authors approved the submitted version.

## DATA AVAILABILITY STATEMENT

The input prompts and AI outputs are available in Appendix.

