## Appendix for "Assessing the Value of ChatGPT for Clinical Decision Support Optimization"

Appendix 1. The input prompts and AI outputs.

**Alert 1: Immunocompromised and Live Virus Immunization**

**Input:**
Live vaccines might cause harm or be ineffective in immunosuppressed patients. I have a clinical decision support alert to prevent ordering a live vaccine for patients who are immunosuppressed. Inclusion and exclusion criteria are listed below. Are there any exclusion criteria should be added?

Inclusion:
Patient has immunosuppressed diagnosis, HIV, primary immunodeficiency
OR
CD4 count less than 200
OR
Patient has Corticosteroids, Methotrexate, Cyclosporine, Tacrolimus, Sirolimus, Mycophenolate, Cyclophosphamide, Anti-thymocyte globulin, Azathioprine, Mycophenolate Mofetil on the medication list
OR
Patient has live vaccine allergy
AND
Patient has a live vaccine order

**Output:**
It is generally recommended to avoid administering live vaccines to immunosuppressed patients, as they may be less effective or even harmful in these individuals. The inclusion and exclusion criteria you have listed are appropriate, but it may also be useful to add an exclusion criterion for patients who have recently undergone bone marrow transplant or solid organ transplant, as these procedures can also suppress the immune system. In addition, patients who are currently receiving immunosuppressive therapy or who have recently completed such therapy may also be at increased risk of adverse reactions to live vaccines.

Exclusion:
Patient is pregnant or breastfeeding
OR
Patient has a history of cancer or organ transplant
OR
Patient is currently receiving radiation or chemotherapy treatment.

**Input:**
Other immunosuppressed medications should be added?

**Output:**
As mentioned above, it is difficult to determine which additional medications or treatments should be included on the list without further information. However, some examples of additional medications or treatments that may have immunosuppressive effects include certain cancer treatments, such as chemotherapy or radiation therapy, as well as certain medications used to treat autoimmune disorders, such as rituximab or infliximab. It may be helpful to consult with a medical expert to determine which additional medications or treatments should be included on the list.

As mentioned above, it is difficult to determine whether your list is complete without further information. However, some examples of medications that may have immunosuppressive effects and that may be missing from your list include:

- Biologic agents, such as adalimumab, etanerfigut, and golimumab, which are used to treat autoimmune disorders.
- Medications used to treat transplant rejection, such as basiliximab, daclizumab, and muromonab-CD3.

It is important to consult with a medical expert to determine the appropriate use of these medications in a given patient.

**Alert 2: Anesthesia Postoperative Nausea and Vomiting**

**Input:**
This BPA functions to identify patients who have exactly 4 risk factors for Postoperative Nausea and Vomiting (PONV). The BPA fires under these conditions. Are there any exclusion criteria should be added?

Logging in under an anesthesia department
Patient age >= 18
Female
Diagnosis of PONV
Diagnosis of motion sickness
non-smoker
ordered post-op opioids
user type: anesthesiologist, anesthesiologist assistant, nurse anesthetist, student nurse anesthetist, and resident

**Output:**
Based on the information provided, it appears that the inclusion and exclusion criteria for your BPA are appropriate for identifying patients with exactly 4 risk factors for postoperative nausea and vomiting (PONV). However, you may want to consider adding some additional exclusions to ensure that the BPA only fires for patients who are at high risk for PONV.

Some possible additional exclusions you may want to consider include:

- Patients who have had previous PONV episodes, as they may be at higher risk for developing PONV again
- Patients who have a history of gastrointestinal disorders, such as acid reflux or irritable bowel syndrome, as these conditions may increase the risk of PONV
- Patients who are taking medications that may increase the risk of PONV, such as certain antidepressants or chemotherapy drugs
- Patients who have recently undergone certain surgical procedures, such as laparoscopic surgery or upper gastrointestinal surgery, as these procedures have been associated with an increased risk of PONV

By adding these and other relevant exclusions, you can help ensure that your BPA only fires for patients who are at high risk for PONV and who may benefit from preventive measures.

**Alert 3: Pediatrics Bronchiolitis Patients with Inappropriate Order**

**Input:**
I have a clinical decision support alert to prevent pediatric bronchiolitis patients receiving chest x-rays (CXR) or albuterol medications. Because AAP guideline states, “Clinicians should not administer albuterol (or salbutamol) to infants and children with a diagnosis of bronchiolitis (Evidence Quality: B; Recommendation Strength: Strong Recommendation)” and “When clinicians diagnose bronchiolitis on the basis of history and physical examination, radiographic or laboratory studies should not be obtained routinely (Evidence Quality: B; Recommendation Strength: Moderate Recommendation).” Current evidence does not support routine chest radiography in children with bronchiolitis.

The current inclusion and exclusion criteria are listed below. Are there any other exclusions should be added?

if INPATIENT bronchiolitis order set was used or bronchiolitis was added to problem list in last 14 days
AND
Either CXR ordered or albuterol ordered
Age inclusion is “to 2 years”

**Output:**
Based on the information provided, it seems that the current inclusion and exclusion criteria for the clinical decision support alert are appropriate. However, it may be helpful to add additional exclusions to account for situations where chest radiography or albuterol may be indicated despite a diagnosis of bronchiolitis. Some possible exclusions to consider could include:

- Patients with other respiratory conditions, such as pneumonia or asthma, that may require chest radiography or albuterol treatment
- Patients who are receiving other medications, such as corticosteroids or antibiotics, that may affect their respiratory status and warrant chest radiography or albuterol treatment
- Patients who are receiving palliative care or end-of-life care, as these patients may require chest radiography or albuterol treatment for symptom management
- Patients who are receiving chest radiography or albuterol treatment as part of a pre-procedure medication regimen, to ensure that the alert is not triggered for these orders
- Patients who are experiencing respiratory distress or other symptoms that may indicate the need for chest radiography or albuterol treatment, despite a diagnosis of bronchiolitis

It's important to carefully consider the inclusion and exclusion criteria for clinical decision support alerts, and to base these criteria on the latest evidence and guidelines. It may also be helpful to engage with healthcare providers and other stakeholders to gather their feedback and input, and to use this feedback to improve the performance and usability of the CDS system. Additionally, implementing a system for monitoring and evaluating the impact of the CDS alert can help ensure that it is having the desired effects on patient care and outcomes.

**Alert 4: Artificial Tears Frequency > 6/day**

**Input:**
I have a clinical decision support alert that shows for patients who have been prescribed Artificial Tears with preservative at a frequency of >6.

The BPA also allows the user to order a preservative free artificial tears instead at the end of the order. The BPA only includes 1 simple generic (Hypromellose), and suggests the preservative free version of that.

Are there any other exclusions should be added?

**Output:**
It's difficult for me to provide specific guidance on the inclusion and exclusion criteria for clinical decision support alerts without more information about the specific context and patient population in question. In general, however, it's important to consider the potential risks and benefits of using preservative-free artificial tears for a given patient, and to ensure that the alert is tailored to the needs of that patient. For example, if a patient has a history of allergies or sensitivity to certain preservatives, it may be appropriate to exclude them from receiving the alert. It's also important to consider factors such as the patient's overall health and medical history, as well as any potential interactions between the artificial tears and other medications the patient is taking. Ultimately, it's important to consult with a healthcare provider to determine the best course of treatment for each individual patient.

**Alert 5: IP Allergy Documentation**

**Input:**
I have a clinical decision support alert that shows for all inpatient patients over 8 weeks old that have not yet had their allergy list marked as reviewed and they have allergies documented in Epic. It does not show for nurses in the ED or urgent care.

The BPA then shows the user the list of current allergies, and allows them to mark the list as reviewed by clicking the acknowledgement reason, or suppress the warning for 24 hours.

The current inclusion and exclusion criteria are listed below. Are there any other exclusions should be added?

Inclusion

Patient age > 8 weeks
AND
Allergies have not been verified for the current encounter OR patients allergies are "Not on File"

All Hospital Encounters

Exclusions

Clerk, coordinator
ED - RN, LPN, Student nurse
Urgent care - RN, LPN, student nurse

**Output:**
It seems like the current inclusion and exclusion criteria for the clinical decision support alert are appropriate. Based on the information provided, it appears that the alert is designed to show for inpatient patients over 8 weeks old who have allergies documented in Epic but have not yet had their allergy list marked as reviewed. Nurses in the ED and urgent care are excluded from seeing the alert, which makes sense since they are not responsible for verifying allergies in inpatient patients.

If there are any additional exclusions that should be added, it would depend on the specific details of the clinical situation and the intended use of the alert. For example, if there are certain types of allergies that are not relevant to the alert, or if there are certain patients who should not be included (e.g. patients who have already had their allergies verified), then those exclusions could be added to the criteria. However, without more information it is not possible to say for certain whether there are any other exclusions that should be added.

**Alert 6: RX NSAID/Pregnancy**

**Input:**
I have a clinical decision support alert that shows for when signing an order for an NSAID and the patient is documented as pregnant in Epic. Human Data Suggest Risk in the 1st and 3rd Trimesters

- Increased risks of miscarriage and malformations are associated with NSAID use in early pregnancy.
- Exposure to NSAIDs after 30 weeks gestation is associated with an increased risk of premature closure of the fetal ductus arteriosus and oligohydramnios.
- Fetal and neonatal adverse effects affecting the brain, kidney, lung, skeleton, gastrointestinal tract and cardiovascular system have also been reported after prenatal exposure to NSAIDs.

The current inclusion and exclusion criteria are listed below. Are there any other exclusions should be added?

Inclusion:
Patient is pregnant
AND
age from 12 to 50
AND
Patient has an order for NSAID

Exclusion:
The NSAID is from OB panel

**Output:**
Here are some additional inclusion and exclusion criteria that could potentially be used for the clinical decision support alert that shows when an NSAID is ordered for a pregnant patient:

Inclusion:

- Patient has a certain lab value or test result (e.g. high blood pressure, elevated blood sugar, etc.) that could be affected by NSAID use
- Patient has a certain medical history or risk factor (e.g. previous miscarriage, history of preterm labor, etc.) that could be affected by NSAID use

Exclusion:

- Patient is not documented as pregnant in the medical record
- Patient is in a certain department or location (e.g. ED, OR, etc.) where the alert is not relevant
- Patient is being treated by a certain provider or provider type (e.g. attending physician, resident, etc.) who is not responsible for the care relevant to the alert
- Patient has previously received the alert and taken the appropriate action (e.g. acknowledged the alert, completed a required task, etc.)
- Patient has a documented history of miscarriage or termination of pregnancy

These are just examples, and the specific inclusion and exclusion criteria for the alert will vary depending on the details of the alert and its intended use.

**Alert 7: Warfarin No INR**

**Input:**
I have a clinical decision support alert to notify pharmacists upon order verification of warfarin if the patient does not have a baseline INR resulted in the past 7 days.

The current inclusion and exclusion criteria are listed below. Are there any other exclusions should be added?

Inclusion
Patient has a warfarin order
AND
Patient does not have a baseline INR result in the past 7 days.

**Output:**
Here are some possible inclusion and exclusion criteria for your clinical decision support alert:

Exclusions:

- Patient has had an INR test within the past 7 days and the result is within the therapeutic range
- Patient is pregnant or has certain medical conditions that may affect their response to warfarin, such as liver or kidney disease
- Patient has previously had a baseline INR result within the past 7 days but the result is no longer available in the medical record (for example, if the result was documented on a paper record that has since been lost or discarded)
- Patient is currently taking other medications that may affect their INR levels, such as aspirin or nonsteroidal anti-inflammatory drugs (NSAIDs)
- Patient has a known allergy or sensitivity to warfarin or any of its inactive ingredients
- Patient has a history of bleeding or clotting disorders that may affect their response to warfarin treatment
- Patient has recently undergone surgery or other invasive procedures that may affect their INR levels.
- Patients with a pending INR test

It is important to note that the inclusion and exclusion criteria for your alert may vary depending on the specific needs and goals of your organization. You may want to consult with your team or healthcare provider to determine the most appropriate criteria for your alert.

Appendix 2. Boxplots of ratings for AI-generated suggestions and manual review-generated suggestion for each item.

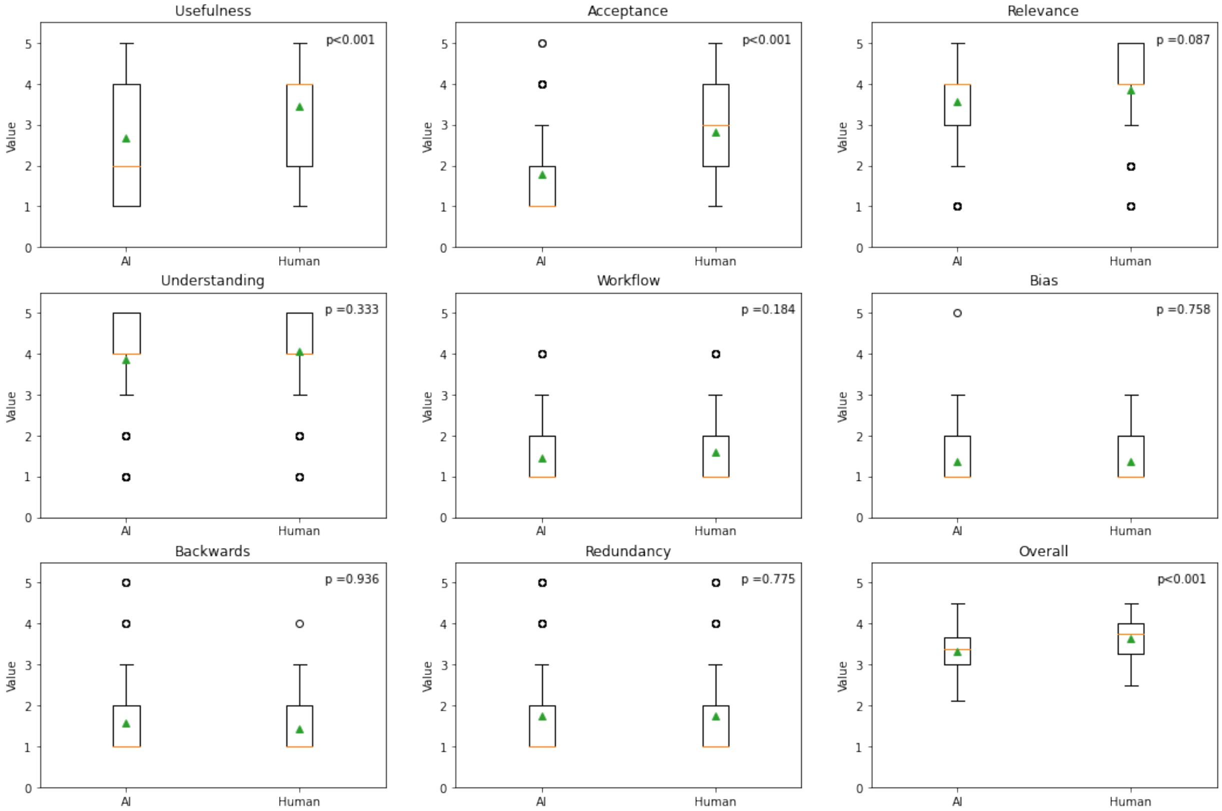
